# HIV Viral Non-Suppression and Its Associated Factors Among PMTCT Mothers Receiving ARV Treatment in Ethiopia

**DOI:** 10.1101/2024.06.13.24308915

**Authors:** Belete Woldesemayat, Ajanaw Yizengaw, Aschale Worku, Amelework Yilma, Sisay Adane, Jaleta Bulti, Eleni Kidane, Gutema Bulti, Saro Abdella

**Affiliations:** HIV AIDS and TB directorate, Ethiopian Public Health Institute, Addis Ababa, Ethiopia; Federal Ministry of Health, Addis Ababa, Ethiopia

**Keywords:** Associated factors, PMTCT, HIV viral non-suppression, Low-level viremia

## Abstract

**Background:** Pediatric HIV infection is mainly caused by Mother-to-child transmission (MTCT). Without any effective medical intervention, 15-45% of infants born to HIV-positive women will become infected with HIV. The contribution of viral non-suppression for MTCT is high. Hence, this study was to determine the magnitude of HIV viral non-suppression, and associated factors among PMTCT Mothers in Ethiopia.

**Methods:** The study was conducted from April 1, 2023, to December 31/ 2024 at 16 public health facilities. An institutional-based cross-sectional study was employed and 496 HIV-positive pregnant and lactating women on ART were included. Data was collected using a prechecked questionnaire with paper mode and ODK (Open Data Kit) for each participant. Venous blood was collected using two Ethylenediamine Tetra acetic Acid (EDTA) tubes for HIV viral load, CD4 and Hgb test. HIV viral load tests were conducted at the EPHI (Ethiopian Public Health Institute) National HIV reference laboratory using the COBAS 4800 System (Roche Molecular Diagnostics). Data was edited and exported to SPSS from the ODK data file, and finally data analysis was performed using SPSS version 26 software.

**Results:** 496 PMTCT Mothers were included in this study, the prevalence of HIV viral non-suppression was 2.2% (95% CI; 1.1-3.9), the rate of viral detectability was 21.8 % (95% CI; 18.2-25.7) and the rate of low-level viremia (LLV) was 9.1%. Family size (X^2^=7.20; p<0.001), a poor and fair level of Adherence (X^2^=18.553; p<0.001), exposure to opportunistic infection (X^2^=25.29; p<0.001), survey time WHO clinical stages II & III (X^2^=25.29; p<0.001), HIV status non-disclosure other than Health care workers (HCWs) (X^2^=4.408; p=0.036), and low survey time CD4 count (<350 cells/ul) ( X^2^= 15.989; p<0.001) were significantly associated to the rate of HIV viral non-suppression.

**Conclusions:** In this study, HIV viral non-suppression among pregnant and lactating women is relatively low (meet the UNAIDS 2030 target). However, Family size, level of Adherence, exposure to opportunistic infection, WHO clinical stage level II and III, HIV status disclosure and low CD4 count were significantly associated with the prevalence of viral non-suppression. To achieve persistent HIV viral suppression enhanced adherence and counselling services should be provided, tailored to the needs of the specific target group.

## 1. Introduction

Mother-to-child transmission (MTCT) of HIV is the primary mode of HIV infection in children and paediatrics [1]. Without any effective medical intervention approximately 15–45% of infants born from HIV-positive women will become infected with HIV, out of these about 5-15% of them are got the infection with breastfeeding [2].

One of the biological risk factors of vertical transmission of HIV is related to the high maternal viral burden, which is a significant predictor of transmission especially when an HIV viral load of more than 1000 Ribonucleic Acid (RNA) copies per one millilitre of plasma [3–4]. In a collaborative prospective study from seven European countries and the United States, MTCT prevalence among anti-retroviral treated women with a viral RNA level of fewer than 1000 copies at close to delivery was only one percent (95% CI; 0.4%–1.9%) [5].

HIV viral load (VL) has a central role in determining MTCT risk, in addition to this; viral lowering or suppression is strongly associated with decreased HIV disease progression, and prevention of sexual transmission. Even if there is a limitation on PMTCT service coverage in developing countries, viral suppression was registered after ART initiation in HIV-infected pregnant women, particularly in sub-Saharan Africa [6]. The year between 2009 and 2014, 89% of infants were prevented with the provision of Anti-retroviral treatment (ART) medication for pregnant and breastfeeding women in 21 high-priority sub-Saharan countries [7].

HIV viral suppression is one of United Nations AIDS’s (UNAIDS’s) targets (95-95-95) to end AIDS epidemics by 2030. The third 95 indicate that at least 95% of those on ART should be virologically suppressed (<1000 RNA copies/ml), translating into a final target of at least 73% and 86% of all PLWH (people living with HIV) achieving virologic suppression [8–9]. It will enable a profound decrease in AIDS-related deaths by 2025 and substantially reduce infection to near-zero levels by 2030 [10]. However, until now there are a lot of impediments, like attitude and behavioural problems towards ART medication, physiological change (e.g. pregnancy) and co-infection with infectious disease (e.g. Syphilis) to achieve the above attainments [11].

On the other hand, different studies showed that because of different factors virological non-suppression and drug resistance are more common in pregnant women than non-pregnant mothers [12–14]. According to the Ethiopian national HIV/AIDS strategic plan, the main way of MTCT is during breastfeeding, which 0.33% of infants were exposed because of the late start of ART during pregnancy, 3.83% of cases were due to dropped off ART from 14,000 spectrum cases [15].

To achieve the optimal protection of MTCT of HIV, early identification (starting from the first antenatal care visit) and during the postnatal period of viral non-suppression during pregnancy and lactating period is very crucial. Many studies have reported on the HIV viral suppression rate and associated factors among non-PMTCT Mothers populations, however, the magnitude of HIV viral non-suppression in this study area and also limited study was reported in our country. Therefore, this study aims to assess the magnitude of viral non-suppression and its associated factors as programs aim for universal access among pregnant and lactating women whose viral load level was non-suppressed in Ethiopia.

## 2. Materials and Methods

### 2.1. Study design and area

An institutional-based cross-sectional study was conducted from April 1/ 2023 to December 31/ 2023 in Addis Ababa, Ethiopia’s capital city, located at 901’48"N 380 44’24"E/9.030000N 98.740000E. According to the Central Statistical Authority of Ethiopia in 2021 population estimation, Addis Ababa has a total population of 3,774,000 people (1,782,000 male and 1,992,000 female), with an annual growth rate of about 2.4%. The city is located in the country’s heartland and is organized into 11 administrative sub-cities (Addissketma, Akaki Kality, Arada, Bole, Gullele, Kirkos, KolfieKeranio, Lideta, Niafas Silk-lafto, Yeka and Lemi-Kura). The health Bureau was in authority of the city’s overall health activities. Numerous clinics, health centres, and hospitals offer medical services [16]. According to Ethiopian Health and Health Related Indicators, there were 78,106 anticipated pregnancies in Addis Ababa and 105,086 delivered by a trained attendant [17].

### 2.1. Study population

All HIV-positive pregnant and lactating mothers, who have been taking ART for at least 6 months and were delivered or come to the health facility for Antenatal care (ANC) and or postnatal care (PNC) services in Sixteen (16) selected health institutions in Addis Ababa and given consent for data and sample collection were recruited in this study. Health facilities (HFs) included in this study have been selected purposively based on their PMTCT client load. We included 5 governmental hospitals, 10 health centres and 01 non-governmental health centre. At the end of the 2022/23 budget year, the total number of PMTCT clients in Addis Ababa was 1041. A total of 16 health facilities were included in this study. Sample size determination was estimated based on a single population proportion by considering different assumptions including, the previous study conducted in West Shewa, Ethiopia, the proportion of viral non-suppression was 11.3% [18] design effect 3, 95% confidence limit and 10% non-respondent rate were considered. The final sample size was 510, and the data were collected from 496 participants in the study period.

### 2.2. Data Collection

Data was collected from 496 study participants by trained health professionals (1 laboratory personnel and 1 nurse/health officer) from ANC and PNC clinics in the selected health facilities. Additional training was given for data collectors on data collection using ODK, laboratory sample collection, sample handling, and storage. Pre-checked questionnaires were filled properly with ODK and paper forms. Sociodemographic (Age, Marital status, Education, Employment status, Family size), clinical and behavioural determinants (Disclosure, Adherence, WHO clinical stage, ART duration, ART regimen, CD4 count, HIV disclosure, Use of memory aids, Use of alternative medicine, Use of alcohol and Substance abuse) and Pregnancy-related variables (Gestational age, Parity, Anemic condition) variables were collected properly. All CD4, Haemoglobin and Syphilis tests were performed during the survey time by the previously existing referral system or in their facility laboratory. After the tests were performed the result was entered and sent by ODK.

Ten (10) millilitre venous blood was collected by using two Ethylene diamine Tetra acetic acid (EDTA) tubes and the plasma sample was separated and prepared immediately within 6 hours of after sample collection according to sample collection and transportation guidelines and stored 2-8 °C. All plasma samples were transported to the EPHI HIV national reference laboratory within 5 days for HIV viral load testing. If the sample was not tested within 5 days after collection, it was stored at −70 °C. Socio-demographic, clinical and other questionnaire-based information was collected by trained nurses or health officers and sample collection, preparation and storage was done by laboratory personnel. A supervisor does daily supervision and sample transportation delegated from EPHI.

### 2.3. Laboratory methods

#### HIV viral load

Viral load testing was conducted at EPHI National HIV Reference Laboratory (NRHIVL) using the Cobas 4800 system (Roche Molecular Diagnostics, Pleasanton CA, USA, 2016 Roche Molecular Systems, Inc.). In clinical specimens, the COBAS Real-Time HIV-1 Quantitative RT-PCR creates the amplified product from the HIV-1 RNA genome, in the process of sample extraction the Cobas 4800 system uses QS (Quantification Standard), an RNA sequence unrelated to the HIV-1 target sequence is added to each specimen. This unrelated RNA sequence is amplified simultaneously with the target sequence by Real-time polymerase chain reaction (RT-PCR) to ensure the sample preparation and amplification procedures are working properly. Using fluorescent-labelled oligonucleotide probes, the COBAS 4800 system analyzes the quantity of HIV-1 target sequence present during each amplification cycle. The probes do not create a signal until they are directly connected to the amplified product. The amplification cycle at which the 4800z detects fluorescence light is proportional to the amount of HIV-1 RNA contained in the original sample. The COBAS 4800 RT system measures the quantity of HIV-1 target sequence present at each amplification cycle using fluorescent-labeled oligonucleotide probes. The fluorescent signal recorded by the PCR machine is proportional to the copies of the HIV-1 RNA concentration in the original sample during the amplification cycle. We used a 0.4 millilitre (ml) sample volume, the lower detection limit is 20 RNA copies per one ml of plasma [19].

### 2.4. Operational definition

#### PMTCT Mothers

Pregnant and lactating women who were taking ARV treatment in the PMTCT clinics

#### Viral non-suppression

≥1000 RNA copies/ml after ART treatment in the plasma sample **Viral Detectability**: Viral load result ≥20 RNA copies/ml in the COBAS 4800 system **Low-level viremia**: Viral load 50 to 999 RNA copies/ml

#### Good Adherence

Participants take medications at prescribed times and frequencies, and follow restrictions regarding food and other medications at least 95%.

#### Poor Adherence

participants take medications at prescribed times and frequencies, and follow restrictions regarding food and other medications less than 95%.

### 2.5. Data processing and analysis

Data was edited and entered into the computer using SPSS software version 26. The summary of descriptive statistics for all dependent and independent variables was done with cross-tabulation and Chi-square analysis. The association between quantitative variables associated with the viral un-suppression was performed using Non-parametric Mann-Whitney U analysis, P values were done and P <0.05 was considered a significant association.

### 2.6. Ethical Approval

Ethical clearance was obtained from the Ethiopian Public Health Institute Scientific Ethical Review Board with the protocol number EPHI-IRB-436-2022. Written informed consent was taken from each study participant. The responders’ personal information was kept private and confidential except for the authors.

## 3. Results

### 3.1. Socio-demographic characteristics of HIV positive Pregnant and Lactating women

In this study 496 participants were included from 16 health facilities; which comprised 5 hospitals, 10 governmental health centres and one non-governmental health centre. The total response rate was 97.3%. Out of 496 participants, 265 (53.4%) were below the age of 30 years, and the mean age of participants was 30.96 ±5.24 (Standard deviation). 153 (30.8%) of participant’s educational status was primary school, 81 (16.3%) had no formal education and 74 (14.9%) of participants’ educational status was college or university. The majority of participants, 427 (86.1%) were Married, 384 (77.4%) were Orthodox Christians, 273 (55%) were housewives and many participants, 285 (57.5%) had family sizes were 3-4 (**Table 1).**

**Table 1:** Socio-demographic characteristics of HIV-positive receiving *ART drugs among* pregnant and lactating women.

| Variables | Category | Frequency | Percent |
| --- | --- | --- | --- |
| Age group | <30 years | 265 | 53.4 |
|  | >=30 years | 231 | 46.6 |
| Educational status | No formal education | 81 | 16.3 |
|  | Primary School | 188 | 37.9 |
|  | Secondary School | 153 | 30.8 |
|  | College/ University | 74 | 14.9 |
| Marital Status | Divorced | 29 | 5.8 |
|  | Widowed | 9 | 1.8 |
|  | Married | 427 | 86.1 |
|  | Single | 31 | 6.3 |
| Religious Belief | Orthodox Christian | 384 | 77.4 |
|  | Protestant | 44 | 8.9 |
|  | Muslim | 62 | 12.5 |
|  | Others | 6 | 1.2 |
| Occupational Status | Government and private employee | 144 | 29.0 |
|  | Housewife | 273 | 55.0 |
|  | Merchant | 31 | 6.3 |
|  | Unemployed | 36 | 7.3 |
|  | Others | 12 | 2.4 |
| Type of health facility | Health Centre | 332 | 66.9 |
|  | Hospital | 164 | 33.1 |
| Family size | 1-2 | 78 | 15.7 |
|  | 3-4 | 285 | 57.5 |
|  | > 4 | 133 | 26.8 |

### 3.2. Clinical and Behavioral Characteristics of Study Participants

Among 496 participants 138 (27.8%) had discordant HIV results, whereas almost half of the participants 244 (49.2%) had the same result (HIV-positive) with their partners. The majority, 455 (91.7%) of participants received regular ART supplies for the last 6 months. Similarly, 465 (93.8%), 461 (92.9%), and 443 (89.3%) participants had good adherence, WHO clinical stage I and had not used any alcohol or other substance for the last month respectively. Out of 496 subjects, 304 (61.3%) had a habit of using memory aid to take ART medicine. On the other hand, among 356 participants who were receiving previous ART regimens the majority, 320 (89.9%) were receiving TDF + 3TC + EFV (**Table 2**).

**Table 2:** Clinical and Behavioural characteristics of PMTCT Mothers.

| Variables | Category | Frequency | Percent |
| --- | --- | --- | --- |
| Partner's HIV status | Negative | 138 | 27.8 |
|  | Positive | 244 | 49.2 |
|  | Unknown | 114 | 23.0 |
| Receiving regular supplies of ART | No | 41 | 8.3 |
|  | Yes | 455 | 91.7 |
| Level of Adherence | Poor | 11 | 2.2 |
|  | Fair | 20 | 4.0 |
|  | Good | 465 | 93.8 |
| Opportunistic infection exposed | No | 461 | 92.9 |
|  | Yes | 35 | 7.1 |
| WHO clinical stage | Stage III | 11 | 2.2 |
|  | Stage II | 24 | 4.8 |
|  | Stage I | 461 | 92.9 |
| Alcohol /substance use in the last one month | No | 443 | 89.3 |
|  | Yes | 53 | 10.7 |
| HIV status disclosure other than HCWs | No | 133 | 26.8 |
|  | Yes | 363 | 73.2 |
| The habit of using a memory aid | No | 192 | 38.7 |
|  | Yes | 304 | 61.3 |
| Previous ART regimen (n=356) | 1B | 2 | 0.6 |
|  | 1C (AZT + 3TC + NVP) | 34 | 9.6 |
|  | 1E (TDF + 3TC + EFV) | 320 | 89.9 |
| ART duration | 6-11 months | 53 | 10.7 |
|  | 1 to 5 years | 175 | 35.3 |
|  | 5 to 10 years | 165 | 33.3 |
|  | more than 10 years | 103 | 20.8 |

In this study for the last 6 months 211 (42.5%) of participants were skipping/stopping ARV medication once or more than once. Out of these 211 participants, 82 (16.5%) skipped/stopped their medication due to they forgot/busy. Similarly, 0.8%, 3.6%, 0.4 %, and 2% skipped/missed their medication because of taking herbal medicines, hospital admission and faith healing respectively (**Figure 1**). According to Figure 2, 455 (91.7%) participants received the new DTG-based ART regimen (TDF, 3TC, DTG). Whereas, 17 (3.4%), 8 (1.6%) and 7 (1.4%) participants were taking ART drugs including, TDF + 3TC + ATV/r, ABC+ 3TC + LPV/r, and TDF + 3TC + EFV respectively. 32 (6.5%) of ART patients were receiving a second-line regimen and the rest 464 (93.5%) were taking a first-line treatment regimen (**Figure 2**).

**Figure 1:**
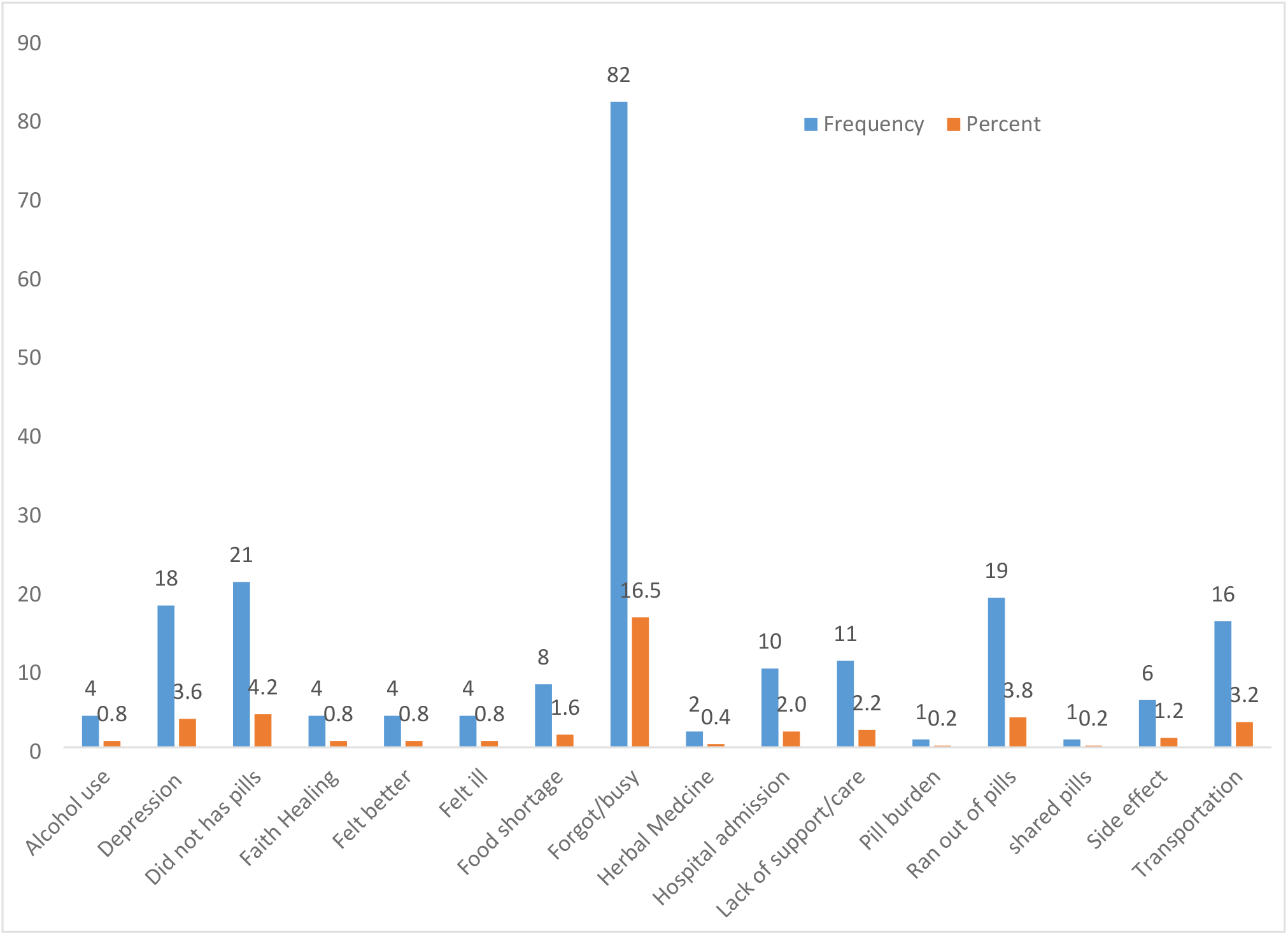
Proportion of participant’s reason to stop/skip ART drugs within the last 6 months (n=211)

**Figure 2:**
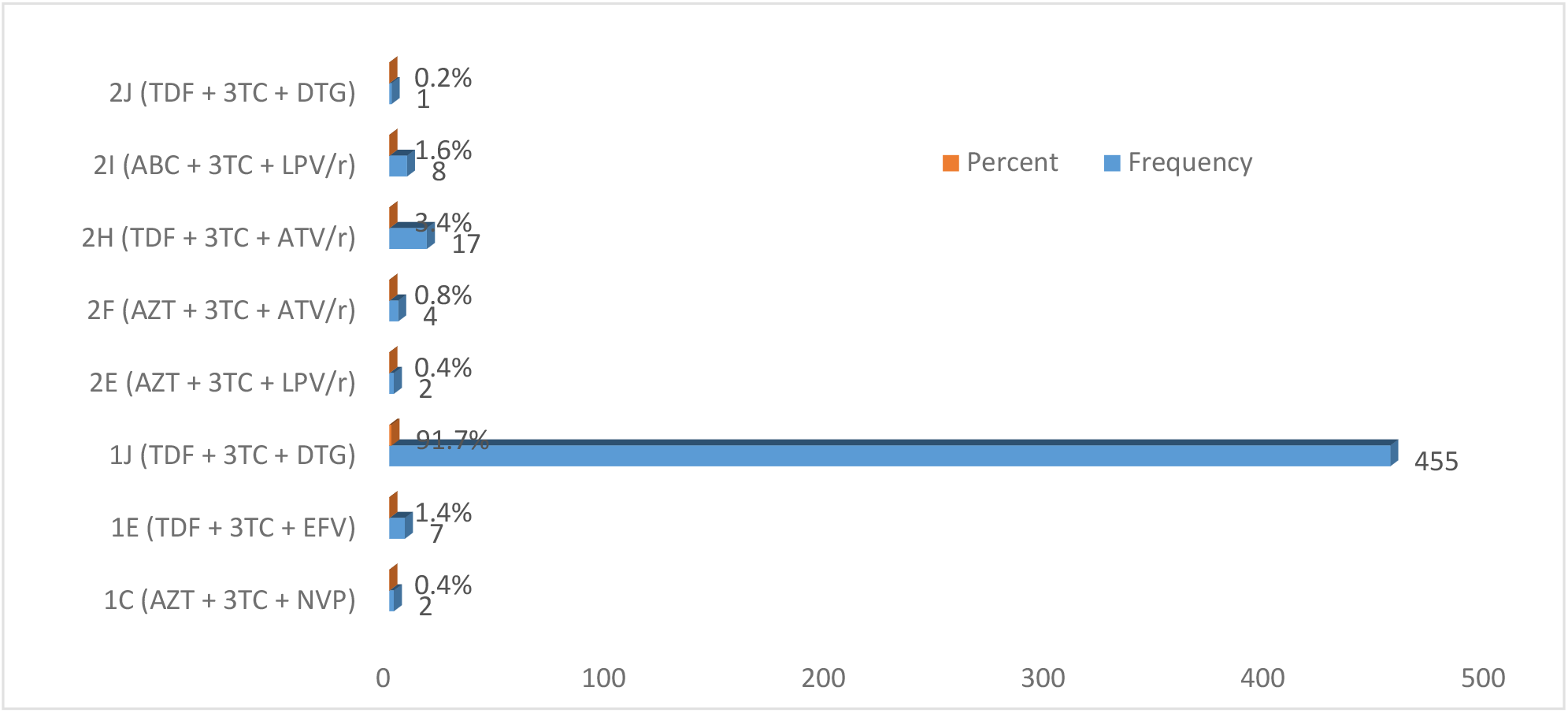
Proportions of ART drug regimen usage among PMTCT Mothers.

According to the obstetrics characteristics of study participants, 320 (64.5%) were lactating women and the rest 176 (35.5%) were HIV-positive pregnant women. Among pregnant women, 11 (6.3%) were in the first trimester and 84 (47.7%) were in the third trimester. 194 (39.1%) of participants had their pregnancy for the first time (primigravida). The majority of participants had 0-1 child (73%), did not have any types of chronic illness (98.2%), had good MUAC measurement (95.4%), ≥ 350 CD4 cells/ul (93.3%) and a negative syphilis antibody test (97.2%) (**Table 3**).

**Table 3:** Obstetrics characteristics of PMTCT Mothers.

| Variables | Category | Frequency | Percent |
| --- | --- | --- | --- |
| Type of participants | Pregnant | 176 | 35.5 |
|  | Lactating | 320 | 64.5 |
| Gestational Age (n=176) | First Trimester | 11 | 6.3 |
|  | Second Trimester | 81 | 46.0 |
|  | Third Trimester | 84 | 47.7 |
| Gravidity | Primigravida | 194 | 39.1 |
|  | Multigravida | 302 | 60.9 |
| Parity | 0-1 child | 362 | 73.0 |
|  | >=2 child | 134 | 27.0 |
| Chronic Illness (at the time of the survey) | No | 487 | 98.2 |
|  | Yes | 9 | 1.8 |
| MUAC | <23 cm | 23 | 4.6 |
|  | >=23 cm | 473 | 95.4 |
| CD4 (time of survey) | <350 cells/ul | 33 | 6.7 |
|  | >=350 | 463 | 93.3 |
| Syphilis Antibody test | Positive | 14 | 2.8 |
|  | Negative | 482 | 97.2 |

### 3.3. Prevalence of HIV-1 viral non-suppression, viral undetectability and low-level viremia

Among 496 HIV-infected pregnant and lactating women who are receiving ART, 11 (2.2%; 95% CI; 1.1 – 3.9) had virologically not suppressed (HIV VL≥1000 copies/ml). The remaining 485 (97.8%) study participants had successfully suppressed viral load (**Figure 3A**). In this study, the magnitude of viral undetectability was 21.8 % (95% CI; 18.2-25.7) (**Figure 3B**). All participants taking first-line or second-line ARV treatment during the survey time were included. The low-level viremia (LLV) (50-999 RNA copies/ml) magnitude of this study was 9.1% (45/496). Participants whose viral load was from 20-50 RNA copies/ml were 52 (10.5%) (**Figure 4**).

**Figure 3:**
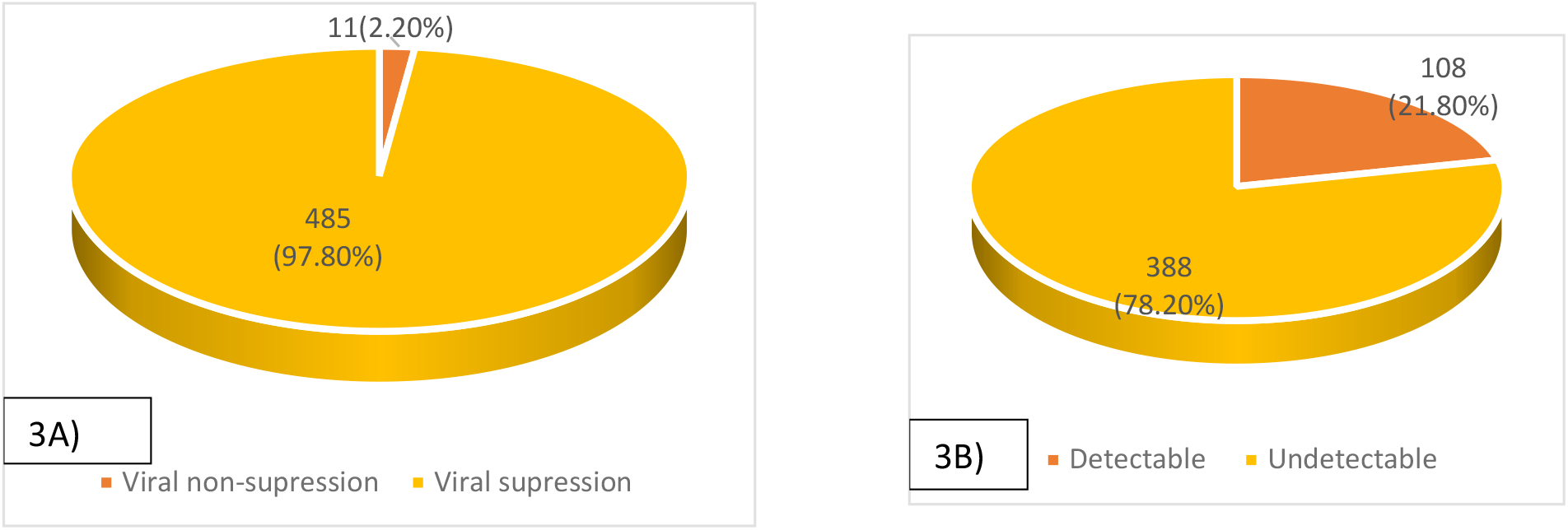
Magnitude of HIV-1 non-suppression and Viral detectability among PMTCT Mothers.

**Figure 4:**
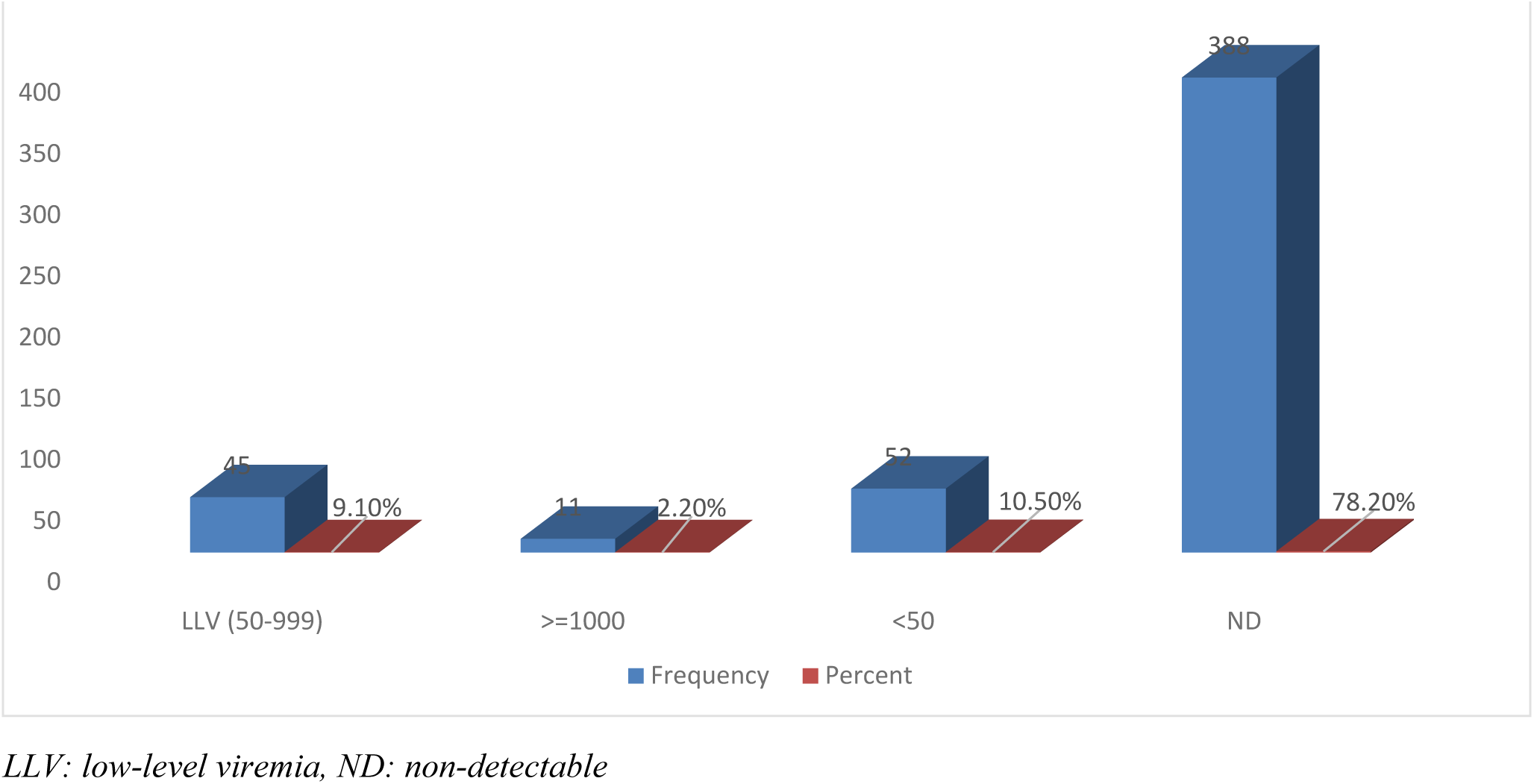
Distribution of low-level viremia, viral non-suppression and viral detectability among PMTCT Mothers.

### 3.4. Associated factors of HIV-1 viral non-suppression

To determine the associated risk factors of HIV-1 viral non-suppression among pregnant and lactating women who were receiving option B+ in the PMTC department in Addis Ababa Health facilities. Socio-demographic, clinical, behavioural, nutritional, obstetric, and immunological determinants were included. To determine the socio-demographic risk factors for the viral non-suppression group among the participants, except family size, all independent variables including; age, marital status, type of health facility, educational status, religious view and occupational status were not significantly associated with non-viral suppression. Regarding family size the proportion of HIV-1 viral non-suppression was high who had a high family size (>4) and according to chi-square analysis the family size is significantly associated with non-viral suppression (X^2^=7.20; P=0.027).

Regarding clinical and behavioural risk factors, participants who had poor adherence and fair adherence were found high proportion of viral non-suppression, poor 1(9.1%) Fair 3(15.0 %), the chi-square analysis showed that Adherence was significantly associated with viral non-suppression (X^2^=18.553; p<0.001). Opportunistic infections during the survey time were significantly associated with viral non-suppression (X^2^=25.290; P<0.001). WHO clinical stage II and III had a high proportion of viral non-suppression (14.3%) compared to WHO clinical stage I (1.3%), the chi-square test also showed that significantly associated with viral non-suppression (X^2^=25.29; P<0.001). Similarly, participants who didn’t disclose their HIV status other than HCWs were one of the associated factors for HIV-1 viral non-suppression (X^2^= 4.408; P=0.036).

The rate of viral non-suppression in patients whose CD4 count was less than 350 cells/ul was 12.1%, whereas the rate of viral non-suppression among those whose CD4 count ≥350 cells/ul in the survey time was 1.5%. Therefore the chi-square analysis showed that CD4 count is significantly associated with HIV-1 viral non-suppression (X^2^=15.989; P<0.001) (**Table 4**).

**Table 4:** *Chi-square* analysis of factors associated with HIV-1 viral non-suppression among PMTCT Mothers.

| Variables | Category | Viral non-suppressed N (%) | Viral suppressed N (%) | Chi-square (X<sup>2</sup>) | P- value |
| --- | --- | --- | --- | --- | --- |
| Type of health facility | Health Centre | 6 (1.8) | 326 (98.2) | 0.780 | 0.377 |
|  | Hospital | 5 (3.0) | 159 (97.0) |  |  |
| Age | <30 years | 4 (1.5) | 261 (98.5) | 1.316 | 0.251 |
|  | >=30 years | 7 (3.0) | 224 (97.0) |  |  |
| Educational status | No formal education | 4 (4.9) | 77 (95.1) | 3.765 | 0.288 |
|  | Primary School | 3 (1.6) | 185 (98.4) |  |  |
|  | Secondary School | 2 (1.3) | 151 (98.7) |  |  |
|  | College/ University | 2 (2.7) | 72 (97.3) |  |  |
| Marital Status | Divorced | 2 (6.9) | 27 (93.1) | 3.510 | 0.319 |
|  | Widowed | 0 (0.0) | 9 (100.0) |  |  |
|  | Married | 8 (1.9) | 419 (98.1) |  |  |
|  | Single | 1(3.2) | 30 (96.8) |  |  |
| Religious Belief | Orthodox Christian | 9 (2.3) | 375 (97.7) | 0.269 | 0.966 |
|  | Protestant | 1 (2.3) | 43 (97.7) |  |  |
|  | Muslim | 1 (1.6) | 61 (98.4) |  |  |
|  | Others | 0 (0.0) | 6 (100.0) |  |  |
| Occupational Status | Government and private employee | 3 (2.1) | 141 (97.9) | 5.409 | 0.248 |
|  | Housewife | 4 (1.5) | 269 (98.5) |  |  |
|  | Merchant | 2 (6.5) | 29 (93.5) |  |  |
|  | Unemployed | 1 (2.8) | 35 (97.2) |  |  |
|  | Others | 1 (8.3) | 11 (91.7) |  |  |
| Family Size | 1-2 | 3 (3.8) | 75 (96.2) | 7.200 | 0.027* |
|  | 3-4 | 2 (0.7) | 283 (99.3) |  |  |
|  | > 4 | 6 (4.5) | 127 (95.5) |  |  |
| Partner's HIV Status | Negative | 5 (3.6) | 133 (96.4) | 4.332 | 0.115 |
|  | Positive | 2 (0.8) | 242 (99.2) |  |  |
|  | Unknown | 4 (3.5) | 110 (96.5) |  |  |
| Receiving regular supplies of ART | No | 2 (4.9) | 39 (95.1) | 1.459 | 0.227 |
|  | Yes | 9 (2.0) | 446 (98.0) |  |  |
| Level of Adherence | Poor | 1 (9.1) | 10 (90.9) | 18.553 | <0.001* |
|  | Fair | 3 (15.0) | 17 (85.0) |  |  |
|  | Good | 7 (1.5) | 458 (98.5) |  |  |
| Opportunistic infection | Yes | 5 (14.3) | 30 (85.7) | 25.290 | <0.001* |
|  | No | 6 (1.3) | 455 (98.7) |  |  |
| WHO clinical stage | Stage II & III | 5 (14.3) | 30 (85.7) | 25.290 | <0.001* |
|  | Stage I | 6 (1.3) | 455 (98.7) |  |  |
| Alcohol/substance use in the last one month | Yes | 3 (5.7) | 50 (94.3) | 3.243 | 0.072 |
|  | No | 8 (1.8) | 435 (98.2) |  |  |
| HIV status disclosure other than HCWs | No | 6 (4.5) | 127 (95.5) | 4.408 | 0.036* |
|  | Yes | 5 (1.4) | 358 (98.6) |  |  |
| The habit of using a memory aid | No | 5 (2.6) | 187 (97.4) | 0.216 | 0.642 |
|  | Yes | 6 (2.0) | 298 (98.0) |  |  |
| Previous ART regimen | 1B | 0 (0.0) | 2 (100.0) | 2.173 | 0.537 |
|  | 1C (AZT + 3TC + NVP) | 0 (0.0) | 34 (100.0) |  |  |
|  | 1E (TDF + 3TC + EFV) | 6 (1.9) | 314 (98.1) |  |  |
| ART duration | 6-11 months | 2 (3.8) | 51 (96.2) | 0.753 | 0.861 |
|  | 1 to 5 years | 4 (2.3) | 171 (97.7) |  |  |
|  | 5 to 10 years | 3 (1.8) | 162 (98.2) |  |  |
|  | more than 10 years | 2 (1.9) | 101 (98.1) |  |  |
| Type of participants | Pregnant | 4 (2.3) | 172 (97.7) | 0.004 | 0.951 |
|  | Lactating | 7 (2.2) | 313 (97.8) |  |  |
| Gestational Age (n=176) | First Trimester | 0 (0.0) | 11 (100.0) | 4.483 | 0.106 |
|  | Second Trimester | 0 (0.0) | 81 (100.0) |  |  |
|  | Third Trimester | 4 (4.8) | 80 (95.2) |  |  |
| Gravidity | Primigravida | 4 (2.1) | 190 (97.9) | 0.036 | 0.850 |
|  | Multigravida | 7 (2.3) | 295 (97.7) |  |  |
| Parity | 0-1 child | 7 (1.9) | 355 (98.1) | 0.499 | 0.480 |
|  | >=2 child | 4 (3.0) | 130 (97.0) |  |  |
| MUAC | <23 cm | 1 (4.3) | 22 (95.7) | 0.505 | 0.477 |
|  | >=23 cm | 10 (2.1) | 463 (97.9) |  |  |
| CD4 | <350 cells/ul | 4 (12.1) | 29 (87.9) | 15.989 | <0.001* |
|  | >=350 | 7 (1.5) | 456 (98.5) |  |  |
| Syphilis Antibody test | Positive | 1 (7.1) | 13 (92.9) | 1.611 | 0.204 |
|  | Negative | 10 (2.1) | 472 (97.9) |  |  |

\*indicates p-value less than <0.05

According to Figure 5, 211 (42.5%) participants skipped/missed their ARV medications at least once in the last 6 months. 6 (3%) of those HIV-1 viral loads were ≥1000 RNA copies/ml, while the rest of the viral suppression among these patients was 97% (n=205). Being forgotten/busy, running out of pills, depression, food shortage, and feeling better have had some associations with the rate of viral non-suppression. However, based on logistic regression analysis the associations were not reached at the significant level (p>0.05) (**Figure 5**).

**Figure 5:**
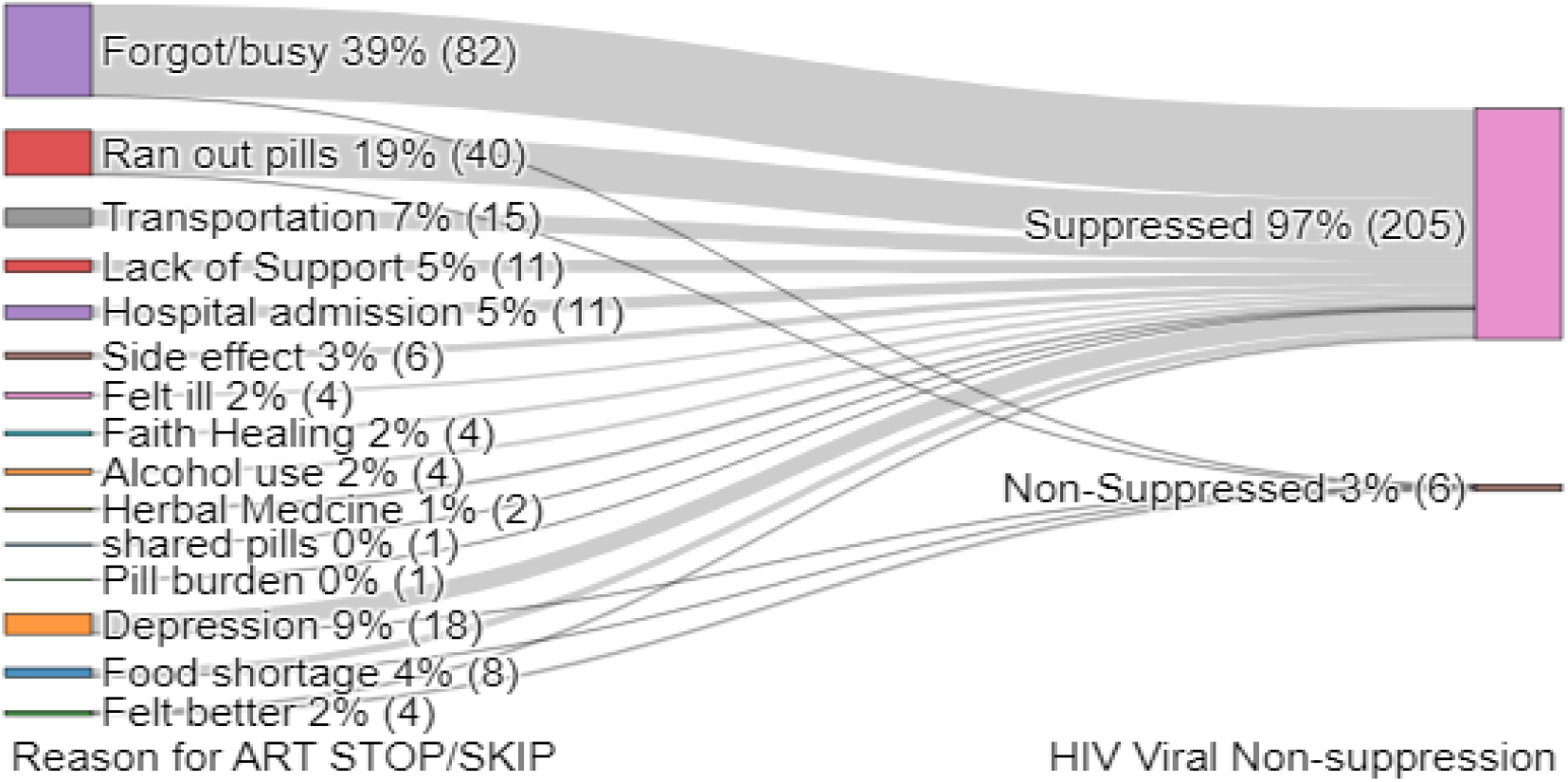
Sunkey diagram of the reason for SKIP/Missed ART drugs and distribution of viral non-suppression among PMTCT Mothers.

According to independent T-test analysis, except for CD4 T cells, all the other parameters (Hgb, MUAC, Age and duration on ART) were not statistically significant association with HIV-1 viral non-suppression. The mean value of CD4 T cell count in the viral non-suppression group was 427 cells/ul and in the suppressed group was 557.85 cells/ul which indicated that the non-suppressed group CD4 is significantly lower compared to the suppressed group (P=0.013)(**Figure 6**). Similarly, the non-parametric Mann-Whitney U analysis showed that only CD4 T cells are significantly associated with HIV-1 Viral non-suppression (**Table 5**).

**Figure 6:**
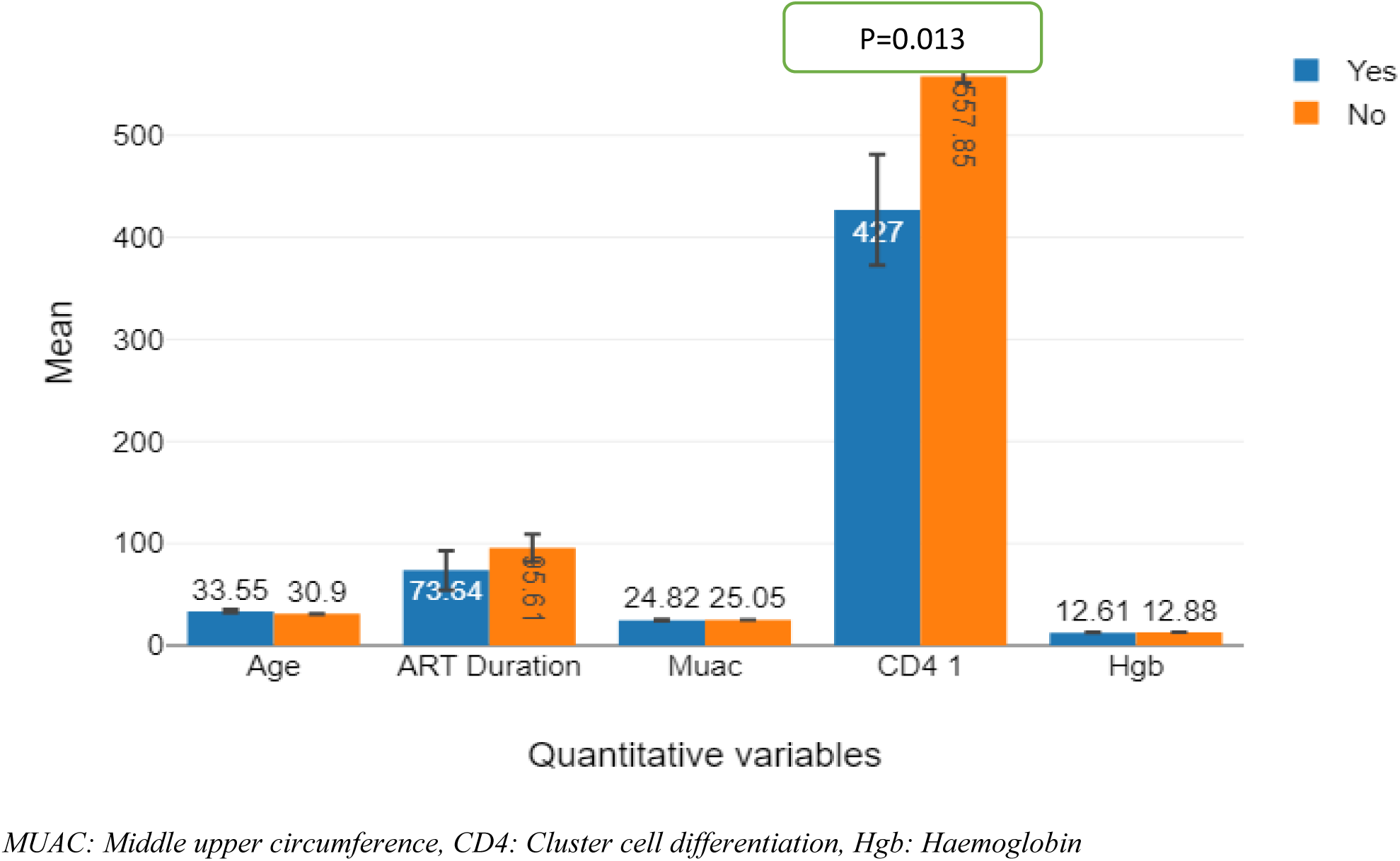
Mean distributions of Quantitative variables in non-suppressed and suppressed PMTCT Mothers.

**Table 5:** Mann-Whitney U analysis of quantitative variables distribution among PMTCT Mothers.

| Quantitative variables | HIV-1 Viral Non-suppressed (N=11) | HIV-1 Viral Suppressed (N=485) | P- value |
| --- | --- | --- | --- |
| Age, Median (IQR) | 35 (30, 37) | 30 (27, 35) | 0.075 |
| Duration on ART in months, Median (IQR) | 60 (15.5, 120) | 72 (28, 120) | 0.696 |
| MUAC measurement in cm, Median (IQR) | 25 (23, 25.53) | 25 (24, 26) | 0.396 |
| CD4 T cell in cells/ul, Median (IQR) | 548 (278, 555) | 555 (542, 563) | 0.013* |
| Heamoglobin in g/dl, Median (IQR) | 12.87 (12.2, 12.92) | 12.88 (12.2, 13.5) | 0.408 |

\*indicates the significance level <0.05

## 4. Discussion

HIV viral non-suppression is a key factor for HIV transmission through vertically or horizontally. The risk of transmission is null to very low if the HIV viral suppression is successful for people who are taking ARV medication. So, to create an AIDS-free generation monitoring viral suppression, enhanced Adherence and counselling in ART drugs, early case detections are the main issues in HIV/AIDS control and prevention programs. Moreover, in HIV-infected women who are taking ARV treatment and could maintain their HIV-1 viral load <1000 RNA copies/ml in pregnancy, delivery and breastfeeding, the mother-to-child HIV transmission could be very low (<%1). Similarly, if their viral load is maintained undetectable throughout their PMTCT period the vertical transmission is almost zero [20]. Our study evaluated the magnitude of HIV-1 Viral non-suppression and associated factors among HIV-positive pregnant and lactating women who are taking ARV treatment for at least six months.

In this study, the prevalence of HIV-1 viral non-suppression was 2.2 % (95% CI; 1.1-3.9), viral detectability was 21.8 % (95% CI; 18.2-25.7) and the magnitude of low-level viremia (LLV) (50-999 RNA copies/ml) was 9.1%. This result is in line with the study conducted in Northern Ethiopia, where the rate of viral non-suppression among pregnant women was 1.31% [21], and a study reported from South Africa, showed that viral non-suppression among women on ART was 4.1% [21]. Similarly, in another study from Northeastern South Africa, the rate of low-level viremia was quite similar to our study (13.8%) [23]. Also, the prevalence of detectable viral load was reported as 17.3 % in Malawi [24]. On the contrary, many previous studies showed that the prevalence of viral non-suppression among pregnant and lactating women was high including in, the Amhara region, Ethiopia (9.1%) [25], Rural South Africa (14.7%) [23], Rural Uganda (19%) [26], Southern and Central Malawi (16%) [27], Mazowe, Zimbabwe (5.6%) [28], Malawi (13.5%) [29], and 12.1% study from Kenya [30]. Our study showed that the prevalence of HIV-1 viral non-suppression is comparatively low, it may mainly be related to the follow-up of PMTCT women increased due to their obstetrics conditions and access to PMTCT service expansion. Additionally, other differences including; type of population, adherence level of the population, geographical and cultural differences and some studies that include ART naïve participants might be the reason for high viral non-suppression. Viral load monitoring programs in the country among PMTCT clients may have a huge impact on viral load suppression, the local health system and program focus could affect the suppression rate [31]. Even though, the prevalence of low viral non-suppression was recorded, it had a critical impact on the program as well as for the offspring since HIV transmission is very high, the recent cohort study result also showed that the risk of HIV transmission was nine times more likely to transmit to the baby if the viral load result is non-suppressed in the time of pregnancy or breastfeeding [32].

Pregnant and lactating mothers whose HIV viral non-suppression was significantly associated with the level of Adherence, presence of opportunistic infection, HIV status disclosure status, family size and CD4 count. These results are supported by many studies which were performed in Ethiopia as well as in different regions of the world. In this study, poor level of adherence and fair level of adherence showed a high proportion of non-viral suppression (14.8%; X^2^=18.55; P= <0.001), a significantly high proportion of non-viral suppression among poor adherence also reported from Amhara region, Ethiopia [25] and Kenya [30]. Exposure to opportunistic infection was one of the associated factors for HIV viral non-suppression in this study, which is in line with the results reported from Uganda [33] and Ethiopia [34], even though both studies were performed on all Adult HIV patients who were taking ART.

The other important factor which is associated with viral non-suppression is the disclosure status of HIV other than health care workers, the proportion of non-disclosure groups is significantly high (4.5%; X^2^=4.41; p=0.036) compared to participants who disclosed those HIV positivity statuses, this result was concordant with the study conducted in Uganda [26] and Kenya [30]. On the other hand, some studies indicated that disclosure status was not significantly associated with the prevalence of non-viral suppression [23, 35]. This difference might be related to cultural differences of the study population, for example, our study population had little or no habit of disclosing their result other than health care workers due to fright of discrimination. It leads to they might not taking their ARV medication to hide themselves from others. In a similar context, this study also revealed that the high numbers of family size (>4 people in the family) was significantly associated (high proportion of viral non-suppression among these women) with viral non-suppression (4.5%; X^2^ = 7.20; p<0.001). The reason for this association might be related to the nutritional status of participants, even if this study showed that the MUAC measurement association did not reach the significant level with the viral non-suppression. In low-income countries high family size is significantly association with poverty and malnutrition [36].

In this study, CD4 count <350 cells/ul in the survey time was significantly associated with the HIV viral non-suppression (12.1 %, X^2^ = 15.989; p<0.001) and the Non-parametric Mann-Whitney U analysis also showed that the viral non-suppression was significantly associated with low CD4 count (p=0.013). This result was concordant with the study in Northern Ethiopia, specifically CD4 count in survey time (< 200 cells/ul) was significantly associated with viral non-suppression [21]. On the other hand, the result reported in Uganda that baseline CD4 count was not associated with non-viral suppression [37]. The difference is related to the time of CD4 cell count because after ART treatment CD4 count could be reversed.

In our study the other factor associated with HIV viral non-suppression was the WHO clinical stage of participants in survey time, the result showed that participants in clinical stage II and Stage III had a statistically significant association with viral non-suppression. 14.3% of participants who were in WHO clinical stages II and III were virally non-suppressed (≥ 1000 RNA copies/ml) ( X^2^= 25.29; p<0.001). In this study, there were no WHO clinical stage IV participants. This result is supported by the study conducted in Ethiopia [25]. According to different studies throughout the World HIV/AIDS disease progression, HIV viral replication and immune suppression (low CD4 count) had closely related outcomes [27, 38]. This study has some limitations including; Since it was an institution-based study we might not have included persons who had poor adherence to ARV drugs due to they had not any follow-up in the data collection period.

## 5. Conclusion

This study revealed that the prevalence of HIV viral non-suppression among pregnant and lactating women is relatively low and it showed that meeting the UNAIDS 2030 target. This performance might be related to the HIV prevention and control program implementation, specifically the PMTCT service improvement from time to time in pregnant and lactating women to prevent HIV transmission risks to their children. However, in this study Family size, level of Adherence, exposure to opportunistic infection, WHO clinical stage level II and III, HIV status disclosure and low CD4 value were significantly associated with the prevalence of viral non-suppression. Even though the prevalence rate was low, enhanced Adherence and counselling should be increased for specific groups of pregnant and lactating women like; women who have large family sizes and do not disclose their HIV status to immediate family members to achieve persistent HIV viral suppression.

## Data Availability

All relevant data are within the manuscript and its Supporting Information files.

NA

## Acknowledgements

Our acknowledgement is forwarded to all staff of the Ethiopian Public Health Institute, the national HIV reference laboratory for valuable support for sample handling and laboratory analysis.

## Author Contributions

**Conceptualization:** Belete Woldesemayat

**Data curation:** Belete Woldesemayat, Ajanaw Yizengaw

**Formal analysis:** Belete Woldesemayat, Ajanaw Yizengaw

**Investigation:** Amelework Yilma, Sisay Adane, Eleni Kidane

**Methodology:** Amelework Yilma, Sisay Adane, Gutema Bulti

**Resources:** Jaleta Bulti, Saro Abdella

**Supervision:** Belete Woldesemayat

**Validation:** Saro Abdella

**Visualization:** Belete Woldesemayat, Ajanaw Yizengaw

**Writing – original draft:** Belete Woldesemayat, Aschale Worku

**Writing–review & editing:** Belete Woldesemayat, Ajanaw Yizengaw, Aschale Worku, Amelework Yilma, Sisay Adane, Eleni Kidane, Gutema Bulti, Jaleta Bulti, Saro Abdella,. All authors have read and agreed to the published version of the manuscript.

